# A viral perspective on worldwide non-pharmaceutical interventions against COVID-19

**DOI:** 10.1101/2020.08.24.20180927

**Authors:** Jean-Philippe Rasigade, Anaïs Barray, Julie Teresa Shapiro, Charlène Coquisart, Yoann Vigouroux, Antonin Bal, Grégory Destras, Philippe Vanhems, Bruno Lina, Laurence Josset, Thierry Wirth

## Abstract

Quantifying the effectiveness of large-scale non-pharmaceutical interventions (NPIs) against COVID-19 is critical to adapting responses against future waves of the pandemic. Most studies of NPIs thus far have relied on epidemiological data. Here, we report the impact of NPIs on the evolution of SARS-CoV-2, taking the perspective of the virus. We examined how variations through time and space of SARS-CoV-2 genomic divergence rates, which reflect variations of the epidemic reproduction number Rt, can be explained by NPIs and combinations thereof. Based on the analysis of 5,198 SARS-CoV-2 genomes from 57 countries along with a detailed chronology of 9 non-pharmaceutical interventions during the early epidemic phase up to May 2020, we find that home containment (35% Rt reduction) and education lockdown (26%) had the strongest predicted effectiveness. To estimate the cumulative effect of NPIs, we modelled the probability of reducing Rt below 1, which is required to stop the epidemic, for various intervention combinations and initial Rt values. In these models, no intervention implemented alone was sufficient to stop the epidemic for Rt’s above 2 and all interventions combined were required for Rt’s above 3. Our approach can help inform decisions on the minimal set of NPIs required to control the epidemic depending on the current Rt value.

---

Coronavirus disease 2019 (COVID-19), caused by the severe acute respiratory syndrome coronavirus 2 (SARS-CoV-2), emerged in China in late 2019^1–3^. Facing or anticipating the pandemic, the governments of most countries implemented a wide range of large-scale non-pharmaceutical interventions, such as lockdown measures, in order to reduce COVID-19 transmission^4–6^. Concerns have been raised regarding the impact of such interventions on the economy, education, and, indirectly, the healthcare system^7^.

Understanding the effectiveness of each non-pharmaceutical intervention against COVID-19 is critical to implementing appropriate responses against current or future waves of the pandemic. Comparative studies of interventions typically rely on epidemiological data to estimate variations of the epidemic reproduction number, which are then correlated with the implementation or relaxation of interventions^5,6,8^. These studies yielded conflicting conclusions. Depending on data sources and epidemiological model design assumptions, some studies identified lockdown (stay at home order) as the most effective intervention^5,9^ while others found little additional impact, if any, compared to other interventions^4,6,10^. Epidemiological studies of interventions against an epidemic face several challenges. Models informed by counts of confirmed cases or deaths ignore the relationships and transmission patterns between cases. Counts themselves can vary in accuracy and timeliness depending on countries’ health facilities, surveillance systems, and the changing definitions of cases. Even when an intervention immediately reduces the transmission rate, a detectable reduction of disease incidence can be much delayed^6^, especially when testing and diagnoses are restricted to specific patient populations. This delay from intervention to incidence reduction, combined with the variety and simultaneous implementation of interventions^4,5^, complicates the identification of their individual effects.

Unlike epidemiological case counts, viral genomes bear phylogenetic information relevant to disease transmission. Extracting this information is the goal of phylodynamics, which relies on evolutionary theory and bioinformatics to model the dynamics of an epidemic^11^. The dates of viral transmission events can also be inferred from genome sequences to alleviate, at least in part, the problems of delayed detection of an intervention’s effect. Here, we conducted a phylodynamic analysis of 5,198 SARS-CoV-2 genomes from 57 countries to estimate the independent effects of 9 large-scale non-pharmaceutical interventions on the transmission rate of COVID-19 during the early dissemination phase of the pandemic. We adapted an established phylogenetic method^12,13^ to model variations of the divergence rate of SARS-CoV-2 in response to interventions and combinations thereof. Building on known relationships between the viral divergence rate and the effective reproduction number *R*_*t*_^14^, we quantified the reduction of *R*_*t*_ independently attributable to each intervention, exploiting heterogeneities in their nature and timing across countries in multivariable models. In turn, these results enabled estimating the probability of stopping the epidemic (*R*_*t*_ < 1) when implementing selected combinations of interventions.

## Survival modelling of viral transmission

The dissemination and detection of a virus in a population can be described as a transmission tree (Fig. 1a) whose shape reflects that of the dated phylogeny of the sampled pathogens (Fig. 1b). In a phylodynamic context, it is assumed that each lineage, represented by a branch in the phylogenetic tree, belongs to a single patient and that lineage divergence events, represented by tree nodes, coincide with transmission events^11^. Thus, branches in a dated phylogeny represent intervals of time between divergence events interpreted as transmission events. This situation can be translated in terms of survival analysis, which models rates of event occurrence, by considering divergence as the event of interest and by treating branch lengths as time-to-event intervals (Fig. 1c, d). Phylogenetic survival analysis was devised by E. Paradis and applied to detecting temporal variations in the divergence rate of tanagers^12^ or fishes^15^, but it has not been applied to pathogens so far^13,16,17^.

**Fig. 1.**
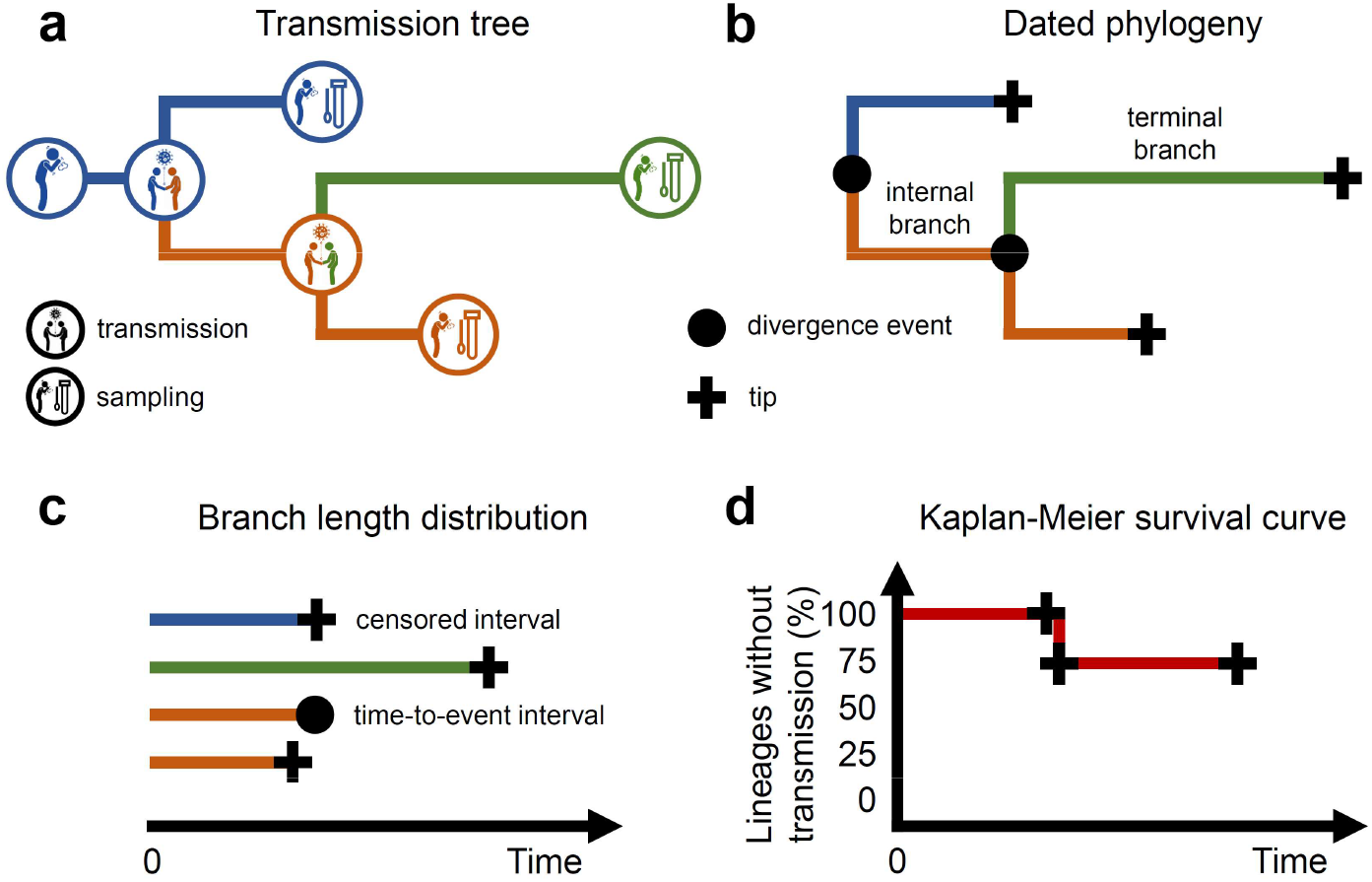
Conceptual overview of phylodynamic survival analysis. Under the assumption that each viral lineage in a phylogeny belongs to an infected patient, the dates of viral transmission and sampling events in a transmission tree (**a**) coincide with the dates of divergence events (nodes) and tips, respectively, of the dated phylogeny reconstructed from the viral genomes (**b**). Treating viral transmission as the event of interest for survival modelling, internal branches connecting two divergence events are interpreted as time-to-event intervals while terminal branches, that do not end with a transmission event, are interpreted as censored intervals (**c**). Translating the dated phylogeny in terms of survival events enables visualizing the probability of transmission through time as a Kaplan-Meier curve (**d**) and modelling the transmission rate using Cox proportional hazards regression.

To quantify the effect of non-pharmaceutical interventions on the transmission rate of COVID-19, we adapted the original model in the Paradis study^12^ to account for the specific setting of viral phylodynamics (see Methods). Hereafter, we refer to the modified model as phylodynamic survival analysis. In survival analysis terms, we interpret internal branches of the phylogeny (those that end with a transmission event) as time-to-event intervals and terminal branches (those that end with a sampling event) as censored intervals (Fig. 1c; see Methods).

The predictors of interest in our setting, namely, the non-pharmaceutical interventions, vary both through time and across lineages depending on their geographic location. To model this, we assigned each divergence event (and subsequent branch) to a country using maximum-likelihood ancestral state reconstruction. Each assigned branch was then associated with the set of non-pharmaceutical interventions that were active or not in the country during the interval spanned by the branch. Intervals containing a change of intervention were split into subintervals^18^. These (sub)intervals were the final observations (statistical units) used in the survival models. Models were adjusted for the hierarchical dependency structure introduced by interval splits and country assignations (see Methods).

## Phylodynamic survival models estimate variations of the reproduction number

The evolution of lineages in a dated viral phylogeny can be described as a birth-death process with a divergence (or birth) rate *λ* and an extinction (or death) rate *μ*^19^. In a phylodynamic context, the effective reproduction number *R*_*t*_ equals the ratio of the divergence and extinction rates^19^. Coefficients of phylodynamic survival models (the so-called hazard ratios of divergence; see Methods) act as multiplicative factors of the divergence rate *λ*, independent of the true value of *λ* which needs not be specified nor evaluated. As *R*_*t*_ = *λ*/*μ*, multiplying *λ* by a coefficient also multiplies *R*_*t*_, independent of the true value of *μ*. Thus, coefficients of phylodynamic survival models estimate variations of *R*_*t*_ in response to predictor variables without requiring external knowledge or assumptions about *λ* and *μ*.

## Variations in COVID-19 transmission rates across countries

We assembled a composite dataset by combining a dated phylogeny of SARS-CoV-2 (Fig. 2a), publicly available from Nextstrain^20^ and built from the GISAID initiative data^21^, with a detailed timeline of non-pharmaceutical interventions available from the Oxford COVID-19 Government Response Tracker (OxCGRT)^22^. Extended Data Fig. 1 shows a flowchart outlining the data sources, sample sizes and selection steps of the study. Phylogenetic and intervention data covered the early phase of the epidemic up to May 4, 2020.

**Figure 2.**
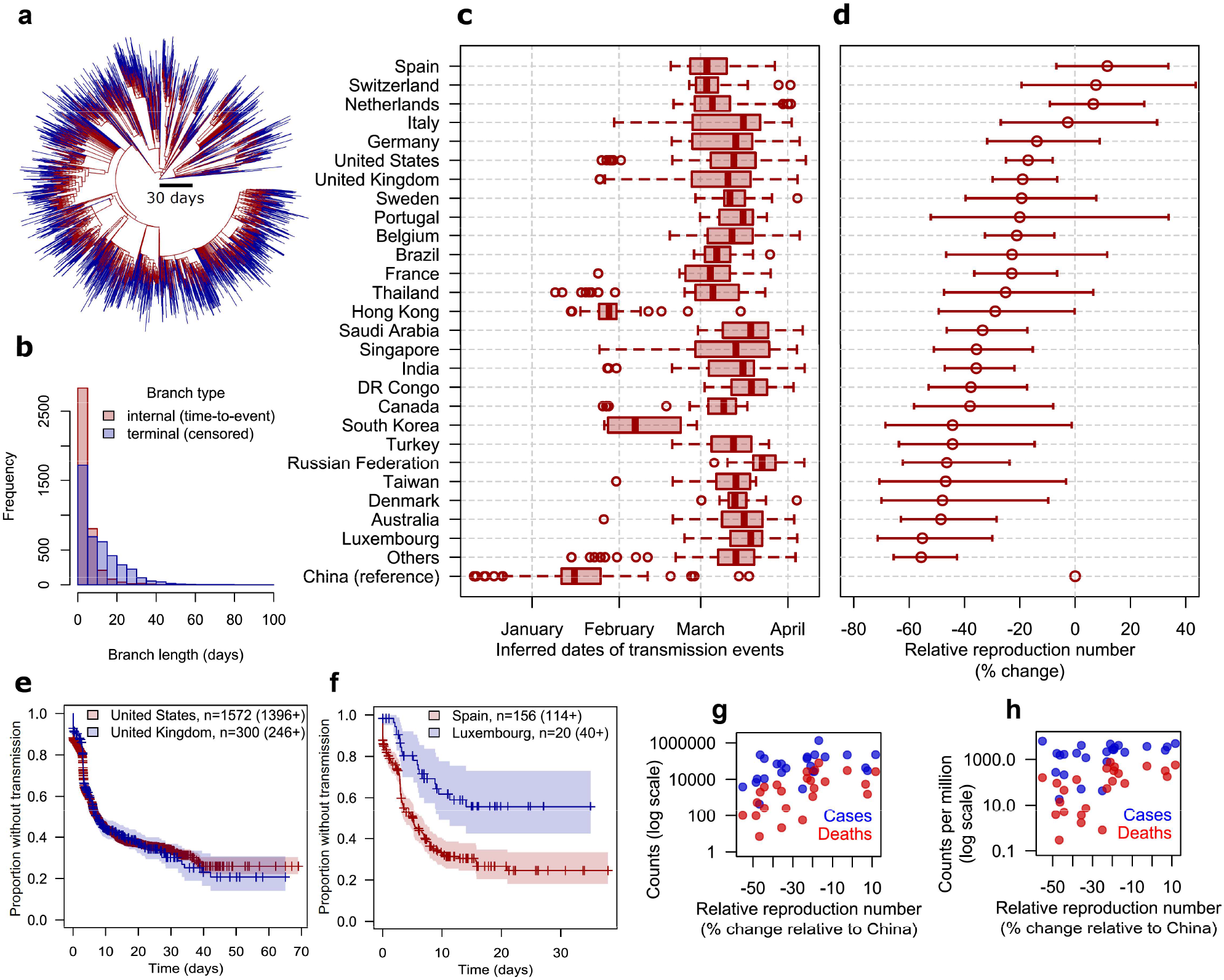
Timing and reproduction numbers of the COVID-19 epidemic in 74 countries based on a dated phylogeny. **a**, Dated phylogeny of 5,198 SARS-CoV-2 genomes where internal (time-to-event) and terminal (time-to-censoring) branches are colored red and blue, respectively. **b**, Histogram of internal and terminal branch lengths. **c**, Box-and-whisker plots of the distribution over time of the inferred transmission events in each country, where boxes denote interquartile range (IQR) and median, whiskers extend to dates at most 1.5x the IQR away from the median date, and circle marks denote dates farther than 1.5 IQR from the median date. **d**, Point estimates and 95% confidence intervals of the relative effective reproduction number, expressed as percent changes relative to China, in 27 countries with ≥10 assigned transmission events. Countries with <10 assigned transmission events (n=32) were pooled into the ‘Others’ category. **e, f**, Representative Kaplan-Meier survival curves of the probability of transmission through time in countries with comparable (**e**) or contrasting (**f**) transmission rates. ‘+’ marks denote censoring events. Numbers denote counts of internal branches and, in brackets, terminal branches. **g, h**, Scatter plots showing correlations between the relative reproduction number and the reported cumulative numbers of COVID-19 cases (blue marks) and deaths (red marks) per country up to May 12, 2020 *(53)*, in absolute values (**g**) and per million inhabitants (**h**).

The 5,198 SARS-CoV-2 genomes used to reconstruct the dated phylogeny were collected from 74 countries. Detailed per-country data including sample sizes are shown in Extended Data Table 1. Among the 10,394 branches in the phylogeny, 2,162 branches (20.8%) could not be assigned to a country with >95% confidence and were excluded, also reducing the number of represented countries from 74 to 59 (Extended Data Fig. 1; a comparison of included and excluded branches is shown in Extended Data Fig. 2). The remaining 4,025 internal branches had a mean time-to-event (delay between transmission events) of 4.4 days (Fig. 2b). These data were congruent with previous estimates of the mean serial interval of COVID-19 ranging from 3.1 days to 7.5 days^23^. The 4,207 terminal branches had a mean time-to-censoring (delay from infection to detection) of 10.6 days (Fig. 2a, b). This pattern of longer terminal vs. internal branches is typical of a viral population in fast expansion^11^.

We compared the timing and dynamics of COVID-19 spread in countries represented in our dataset (Fig. 2c, d), pooling countries with <10 assigned transmission events into an ‘others’ category. The estimated date of the first local transmission event in each country showed good concordance with the reported dates of the epidemic onset (Pearson correlation = 0.84; Extended Data Fig. 3). The relative effective reproduction number *R*_*t*_ per country, taking China as reference, ranged from - 55.6% (95%CI, −71.4% to −29.9%) in Luxembourg to +11.7% (95% CI, −6.7% to +33.8%) in Spain (Fig. 2c). Notice that these estimates are averages over variations of *R*_*t*_ through time in each country, up to May 4, 2020. Exemplary survival curves of transmission events are shown in Fig. 2e, f. Relative *R*_*t*_’s are not expected to necessarily correlate with the reported counts of COVID-19 cases across all countries due to the confounding effects of population sizes, case detection policies and the number of genomes included. Nevertheless, the relative *R*_*t*_’s across countries substantially correlated with the reported cumulative counts up to May 12 (Fig. 2g, h), including COVID-19 cases (Pearson correlation with log-transformed counts, 0.46, 95% CI, 0.07 to 0.73), deaths (correlation 0.59, 95% CI, 0.24 to 0.80), cases per million inhabitants (correlation 0.39, 95% CI, −0.01 to 0.69) and deaths per million inhabitants (correlation 0.56, 95% CI, 0.21 to 0.79).

## Disentangling the individual effectiveness of non-pharmaceutical interventions

The implementation and release dates of large-scale non-pharmaceutical interventions against COVID-19 were available for 57 countries out of the 59 represented in the dated phylogeny. Definitions of the selected interventions are shown in Table 1. Branches assigned to countries with missing intervention data, namely, Latvia and Senegal, were excluded from further analysis (n=22/8,262 (0.3%); see Extended Data Fig. 1). Up to May 4, 2020, the interventions most universally implemented were information campaigns, restrictions on international travel and education lockdown (>95% of countries) (Extended Data Fig. 4). The least frequent were the closure of public transportation (35%) and home containment (72%). Public information campaigns came first and home containment came last (median delay across countries, 5 days before and 24 days after the first local transmission event, respectively; Fig. 3a). Survival curves for each intervention are shown in Extended Data Fig. 5. Most interventions were implemented in combination and accumulated over time rather than replacing each other (Extended Data Fig. 4; median delays between interventions are shown in Extended Data Fig. 6; correlations in Extended Data Fig. 7; and a detailed timeline of interventions in Extended Data Table 2). However, we observed a substantial heterogeneity of intervention timing across the 57 countries (Fig. 3a), suggesting that individual intervention effects can be discriminated by multivariable analysis given the large sample size (n=8,210 subintervals).

**Table 1.**
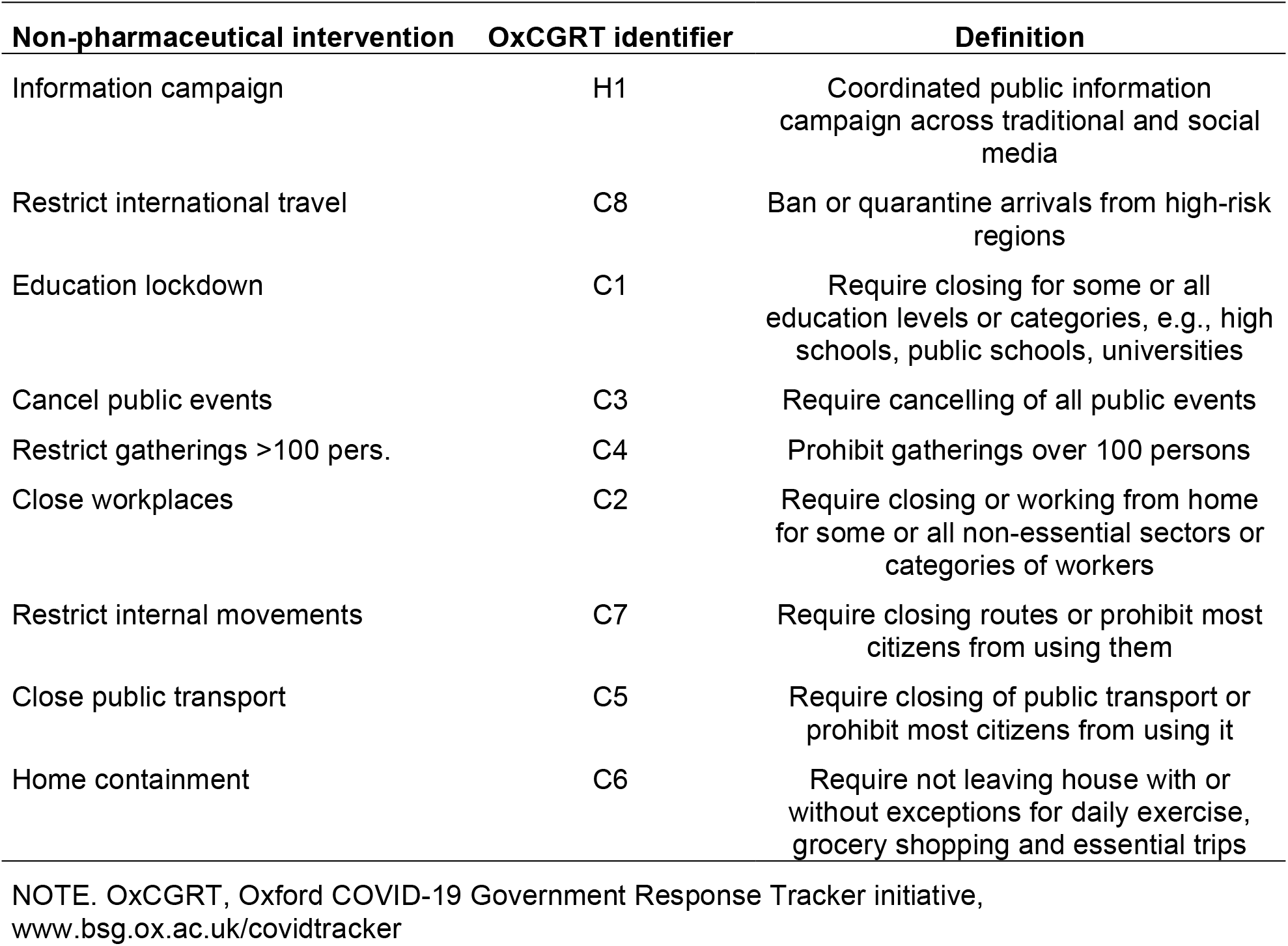
Selected large-scale non-pharmaceutical interventions against COVID-19.

**Figure 3.**
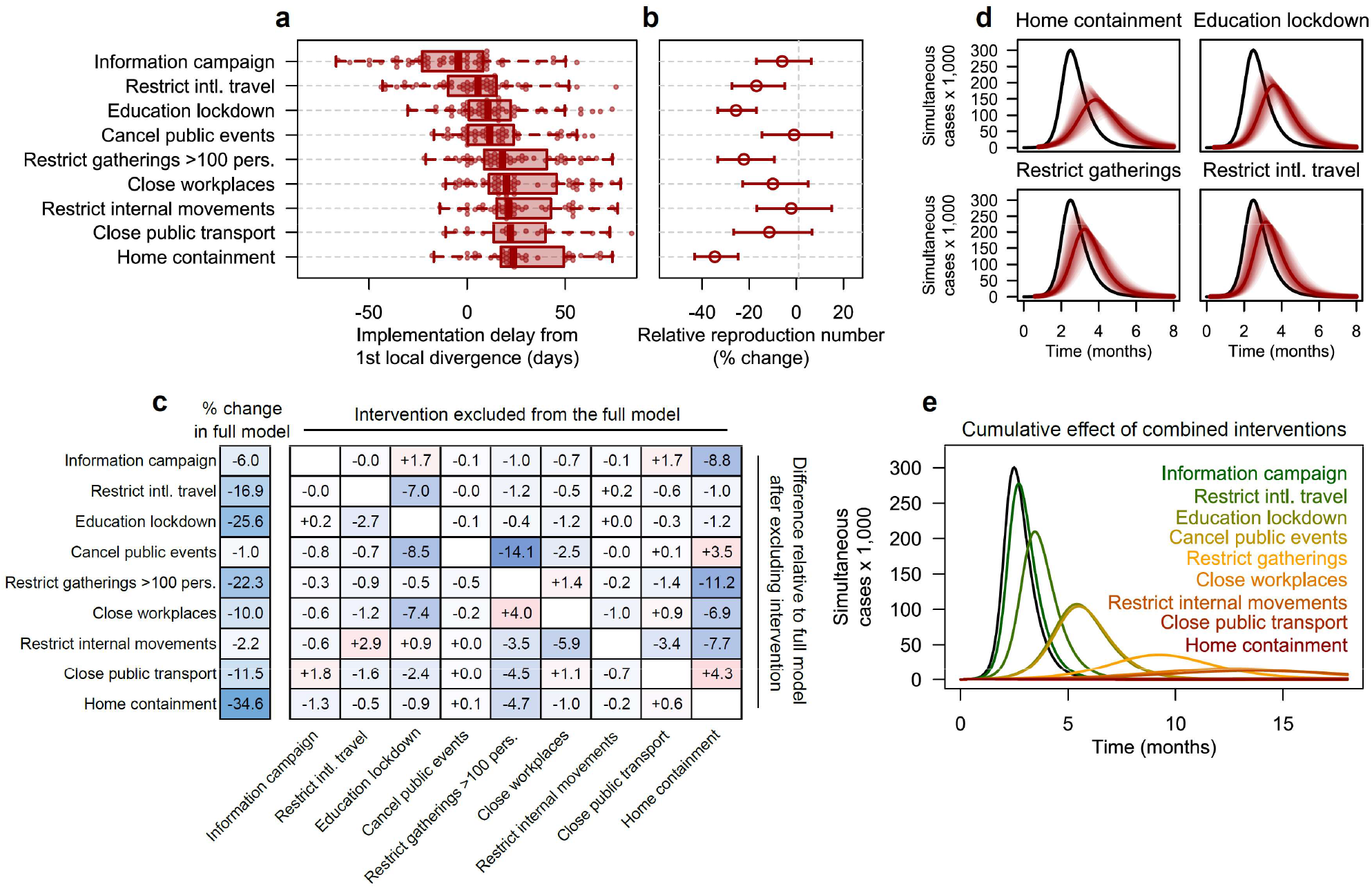
Non-pharmaceutical interventions variably reduce the reproduction number of COVID-19. Data derive from the phylogenetic survival analysis of 4,191 internal and 4,019 terminal branches of a dated phylogeny of SARS-CoV-2 genomes, combined with a chronology of interventions in 57 countries. **a**, Box-and-whisker plots of the delay between the 1st SARS-CoV-2 divergence event and the intervention. Plot interpretation is similar with Fig. 2c. **b**, Point estimates and 95% confidence intervals of the independent % change of the effective reproduction number predicted by each intervention in a multivariable, mixed-effect phylogenetic survival model adjusted for between-country variations. **c**, Matrix of pairwise interactions between the interventions (in rows) estimated using 9 multivariable models (in columns), where each model ignores exactly one intervention. Negative (positive) differences in blue (red) denote a stronger (lesser) predicted effect of the intervention in row when ignoring the intervention in column. **d, e**, Simulated impact of interventions implemented independently (**d**) or in sequential combination (**e**) on the count of simultaneous cases in an idealized population of 1 million susceptible individuals using compartmental SIR models with a basic reproduction number *R*_0_ = 3 (black lines) and a mean infectious period of 2 weeks. Shaded areas in (**d**) denote 95% confidence bands.

A multivariable phylogenetic survival model, including the 9 interventions and controlling for between-country *R*_*t*_ variations (see Methods), showed a strong fit to the data (likelihood-ratio test compared to the null model, P < 10^−196^). In this model, the interventions most strongly and independently associated with a reduction of the effective reproduction number *R*_*t*_ of SARS-CoV-2 were home containment (*R*_*t*_ percent change, −34.6%, 95%CI, −43.2 to −24.7%), education lockdown (−25.6%, 95%CI, −33.4 to −16.9%), restricting gatherings (−22.3%, 95%CI, −33.4 to −9.4%) and international travel (−16.9%, 95%CI, −27.5 to −4.8%). We failed to detect a substantial impact of other interventions, namely information campaigns, cancelling public events, closing workplaces, restricting internal movement, and closing public transportation (Fig. 3b). Based on coefficient estimates, all interventions were independently predicted to reduce *R*_*t*_ (even by a negligible amount), in line with the intuition that no intervention should accelerate the epidemic. Contrasting with previous approaches that constrained coefficients^5^, this intuition was not enforced a priori in our multivariable model, in which positive coefficients (increasing *R*_*t*_) might have arisen due to noise or collinearity between interventions. The absence of unexpectedly positive coefficients suggests that our model correctly captured the epidemic slowdown that accompanied the accumulation of interventions.

## Estimated intervention effects are robust to time-dependent confounders and collinearity

A reduction of *R*_*t*_ through time, independent of the implementation of interventions, might lead to overestimate their effect in our model. Several potential confounders might reduce *R*_*t*_ through time but cannot be precisely estimated and included as control covariates. These included the progressive acquisition of herd immunity, the so-called artificial diversification slowdown possibly caused by incomplete sampling, and time-dependent variations of the sampling effort (see Methods). To quantify this potential time-dependent bias, we constructed an additional model including the age of each branch as a covariate (Extended Data Table 1). The coefficients in this time-adjusted model only differed by small amounts compared to the base model. Moreover, the ranking by effectiveness of the major interventions remained unchanged, indicating that our estimates were robust to time-dependent confounders.

We also quantified the sensitivity of the estimated intervention effects to the inclusion of other interventions (collinearity) by excluding interventions one by one in 9 additional models (Fig. 3c). This pairwise interaction analysis confirmed that most of the estimated effects were strongly independent. Residual interferences were found for gathering restrictions, whose full-model effect of −22.3% was reinforced to −33.5% when ignoring home containment; and for cancelling public events, whose full-model effect of −0.97% was reinforced to −15.1% when ignoring gathering restrictions. These residual interferences make epidemiological sense because home containment prevents gatherings and gathering restrictions also prohibit public events. Overall, the absence of strong interferences indicated that our multivariable model reasonably captured the independent, cumulative effect of interventions, enabling ranking their impact on COVID-19 spread.

## Simulating intervention effectiveness in an idealized population

To facilitate the interpretation of our estimates of the effectiveness of interventions against COVID-19, we simulated each intervention’s impact on the peak number of cases, whose reduction is critical to prevent overwhelming the healthcare system (Fig. 3d and Extended Data Fig. 8). We used compartmental Susceptible-Infected-Recovered (SIR) models with a basic reproduction number *R*_0_ = 3 and a mean infectious period of 2 weeks based on previous estimates^24,25^, in an idealized population of 1 million susceptible individuals (see Methods). In each model, we simulated the implementation of a single intervention at a date chosen to reflect the median delay across countries (Fig. 3a) relative to the epidemic onset (see Methods). On implementation date, the effective reproduction number was immediately reduced according to the estimated intervention’s effect shown in Fig. 3b.

In this idealized setting, home containment, independent of all other restrictions, only halved the peak number of cases from 3.0×10^5^ to 1.5×10^5^ (95% CI, 1.0×10^5^ to 2.0×10^5^) (Fig. 3d). However, a realistic implementation of home containment also implies other restrictions including, at least, restrictions on movements, gatherings, and public events. This combination resulted in a relative *R*_*t*_ of −50.8% (95% CI, −59.4% to - 40.2%) and a 5-fold reduction of the peak number of cases to 6.0×10^4^ (95% CI, 1.9×10^4^ to 1. ×10^5^). Nevertheless, if *R*_0_ = 3 then a 50% reduction is still insufficient to reduce *R*_*t*_ below 1 and stop the epidemic. This suggests that even when considering the most stringent interventions, combinations may be required. To further examine this issue, we estimated the effect of accumulating interventions by their average chronological order shown in Fig. 3a, from information campaigns alone to all interventions combined including home containment (Fig. 3e). Strikingly, only the combination of all interventions completely stopped the epidemic under our assumed value of *R*_0_. To estimate the effectiveness of combined interventions in other epidemic settings, we computed the probabilities of reducing *R*_*t*_ below 1 for values of *R*_0_ ranging from 1.5 to 3.5 (Table 2; see Methods). The same probabilities for individual interventions are presented in Table S2, showing that no single intervention would stop the epidemic if *R*_0_ ≥ 2. These results may help inform decisions on the appropriate stringency of the restrictions required to control the epidemic under varying transmission regimes.

**Table 2.** Predicted reduction of the COVID-19 effective reproduction number under increasingly stringent combinations of non-pharmaceutical interventions.

| Accumulated interventions | Relative $R_t$<br>(cumulative % change) | Probability of reducing $R_t$ below 1 | | | | |
| --- | --- | --- | --- | --- | --- | --- |
| | | $R_0=1.5$ | $R_0=2.0$ | $R_0=2.5$ | $R_0=3.0$ | $R_0=3.5$ |
| Information campaign | -6.0 (-17.0 to +6.5) | <0.01 | <0.01 | <0.01 | <0.01 | <0.01 |
| + Restrict intl. travel | -21.9 (-35.0 to -6.1) | 0.05 | <0.01 | <0.01 | <0.01 | <0.01 |
| + Education lockdown | -41.9 (-52.5 to -29.0) | 0.91 | 0.07 | <0.01 | <0.01 | <0.01 |
| + Cancel public events | -42.5 (-54.0 to -28.1) | 0.90 | 0.11 | <0.01 | <0.01 | <0.01 |
| + Restrict gatherings >100 pers. | -55.3 (-63.4 to -45.5) | 1.00 | 0.86 | 0.14 | <0.01 | <0.01 |
| + Close workplaces | -59.7 (-67.6 to -50.0) | 1.00 | 0.98 | 0.48 | 0.04 | <0.01 |
| + Restrict internal movements | -60.6 (-67.9 to -51.6) | 1.00 | 0.99 | 0.56 | 0.06 | <0.01 |
| + Close public transport | -65.1 (-72.6 to -55.7) | 1.00 | 1.00 | 0.87 | 0.36 | 0.05 |
| + Home containment | -77.2 (-81.5 to -71.9) | 1.00 | 1.00 | 1.00 | 1.00 | 0.98 |

## Discussion

We present a phylodynamic analysis of how the divergence rate and reproduction number of SARS-CoV-2 varies in response to large-scale non-pharmaceutical interventions in 57 countries. Our results suggest that no single intervention, including home containment, is sufficient on its own to stop the epidemic (*R*_*t*_ < 1). Increasingly stringent combinations of interventions may be required depending on the effective reproduction number.

Home containment was repeatedly estimated to be the most effective response in epidemiological studies from China^26^, France^27^, the UK^28^, and Europe^5^. Other studies modelled the additional (or residual) reduction of *R*_*t*_ by an intervention after taking into account those previously implemented^4,6,10^. Possibly because home containment was the last implemented intervention in many countries (Fig. 2a), these studies reported a weaker or even negligible additional effect compared to earlier interventions. In our study, home containment, even when implemented last, had the strongest independent impact on epidemic spread (*R*_*t*_ percent change, −34.6%), which was further amplified (−50.8%) when taking into account implicit restrictions on movements, gatherings and public events.

We found that education lockdown substantially decreased COVID-19 spread (*R*_*t*_ percent change, −25.6%). Contrasting with home containment, the effectiveness of education lockdown has been more hotly debated. This intervention ranked among the most effective ones in two international studies^4,6^ but had virtually no effect on transmission in other reports from Europe^5,10^. Young children have been estimated to be poor spreaders of COVID-19 and less susceptible than adults to develop disease after an infectious contact, counteracting the effect of their higher contact rate^29,30^. However, the relative susceptibility to infection was shown to increase sharply between 15 and 25 years, suggesting that older students might be more involved in epidemic spread^30^. Importantly, we could not differentiate the effect of closing schools and universities because both closures coincided in all countries. Thus, our finding that education lockdowns reduce COVID-19 transmission might be driven by contact rate reductions in older students rather than in children, as hypothesized elsewhere^4^. This raises the question of whether education lockdown should be adapted to age groups, considering that: (1), education lockdown had a sizeable impact on COVID-19 transmission in our study and others^4,6^; (2), this impact might be preferentially driven by older students^29,30^; and (3) autonomous distance learning might be more effective in university students^31^ compared to younger children who require parental presence and supervision following school closure, possibly widening the gap for children from under-resourced backgrounds^32^. Based on these elements, we speculate that closing universities, but not elementary schools, might strike the right balance between efficacy and social impact.

Restrictions on gatherings of >100 persons appeared more effective than cancelling public events (*R*_*t*_ percent changes, −22.3% vs. −1.0%, respectively) in our phylodynamic model, in line with previous results from epidemiological models^4^. Notwithstanding that gathering restrictions prohibit public events, possibly causing interferences between estimates (Fig. 3c), this finding is intriguing. Indeed, several public events resulted in large case clusters, the so-called superspreading events, that triggered epidemic bursts in France^33^, South Korea^34^ or the U.S.^35^. A plausible explanation for not detecting the effectiveness of cancelling public events is that data-driven models, including ours, better capture the cumulative effect of more frequent events such as gatherings than the massive effect of much rarer events such as superspreading public events. This bias towards ignoring the so-called ‘Black Swan’ exceptional events^36^ suggests that our findings (and others’^4^) regarding restrictions on public events should not be interpreted as an encouragement to relax these restrictions but as a potential limit of modelling approaches (but see^37^).

There are other limitations to our study, including its retrospective design. Similar to previous work^6^, we did not consider targeted non-pharmaceutical interventions that are difficult to date and quantify, such as contact tracing or case isolation policies. Data were analyzed at the national level, although much virus transmission was often concentrated in specific areas and some non-pharmaceutical interventions were implemented at the sub-national level^38^. Our phylogeographic inferences did not consider the travel history of patients, whose inclusion in Bayesian models was recently shown to alleviate sampling bias^39^. From a statistical standpoint, the interval lengths in the dated phylogeny were treated as fixed quantities in the survival models. Ignoring the uncertainty of the estimated lengths might underestimate the width of confidence intervals, although this is unlikely to have biased the pointwise estimates and the ranking of interventions’ effects. The number of genomes included by country did not necessarily reflect the true number of cases, which might have influenced country comparison results in Fig. 2, but not intervention effectiveness models in Fig. 3 which were adjusted for between-country variations of *R*_*t*_. Finally, our estimates represent averages over many countries with different epidemiological contexts, healthcare systems, cultural behaviors and nuances in intervention implementation details and population compliance. This global approach, similar to previous work^4,6^, facilitates unifying the interpretation of intervention effectiveness, but this interpretation still needs to be adjusted to local contexts by policy makers.

Beyond the insights gained into the impact of interventions against COVID-19, our findings highlight how phylodynamic survival analysis can help leverage pathogen sequence data to estimate epidemiological parameters. Contrasting with the Bayesian approaches adopted by most, if not all, previous assessments of intervention effectiveness^4,5,8^, phylodynamic survival analysis does not require any quantitative prior assumption or constraint on model parameters. The method should also be simple to implement and extend by leveraging the extensive software arsenal of survival modelling. Phylodynamic survival analysis may complement epidemiological models as pathogen sequences accumulate, allowing to address increasingly complex questions relevant to public health strategies.

## METHODS

### Definitions and chronology of non-pharmaceutical interventions

The nature, stringency and timing of non-pharmaceutical interventions against COVID-19 have been collected and aggregated daily since January 1, 2020 by the Oxford COVID-19 Government Response Tracker initiative of the Blavatnik School of Government, UK^4,22^. As of May 12, 2020, the interventions are grouped into three categories, namely: closures and containment (8 indicators), economic measures (4 indicators) and health measures (5 indicators). Indicators use 2-to 4-level ordinal scales to represent each intervention’s stringency, and an additional flag indicating whether the intervention is localized or general. Details of the coding methods for indicators can be found in^40^. We focused on large-scale interventions against transmission that did not target specific patients (for instance, we did not consider contact tracing) and we excluded economic and health interventions except for information campaigns. This rationale led to the selection of the 9 indicators shown in Table 1. To facilitate interpretation while constraining model complexity, the ordinal-scale indicators in OxCGRT data were recoded as binary variables in which we only considered government requirements (as opposed to recommendations) where applicable. We did not distinguish between localized and nation-wide interventions because localized interventions, especially in larger countries, targeted the identified epidemic hotspots. As the data did not allow to differentiate closures of schools and universities, we use the term ‘education lockdown’ (as opposed to ‘school closure’ in^22^) to avoid misinterpretation regarding the education levels concerned.

### Phylodynamic survival analysis in measurably evolving populations

The original phylogenetic survival model in^12^ and its later extensions^41^ considered intervals backward in time, from the tips to the root of the tree, and were restricted to trees with all tips sampled at the same date relative to the root (ultrametric trees). Censored intervals (intervals that do not end with an event) in^12^ were used to represent lineages with known sampling date but unknown age. In contrast, viral samples in ongoing epidemics such as COVID-19 are typically collected through time. A significant evolution of the viruses during the sampling period violates the ultrametric assumption. To handle phylogenies of these so-called measurably evolving populations^42^, we propose a different interpretation of censoring compared to^12^. Going forward in time, the internal branches of a tree connect two divergence events while terminal branches, those that end with a tip, connect a divergence event and a sampling event (Fig. 1b). Thus, we considered internal branches as time-to-event intervals and terminal branches as censored intervals representing the minimal duration during which no divergence occurred (Fig. 1c).

### SARS-CoV-2 phylogenetic data

SARS-CoV-2 genome sequences have been continuously submitted to the Global Initiative on Sharing All Influenza Data (GISAID) by laboratories worldwide^21^. To circumvent the computational limits of phylogeny reconstruction and time calibration techniques, the sequences of the GISAID database are subsampled before analysis by the Nextstrain initiative, using a balanced subsampling scheme through time and space^20,43^. Phylogenetic reconstruction uses maximum-likelihood phylogenetic inference based on IQ-TREE^44^ and time-calibration uses TreeTime^45^. See^46^ for further details on the Nextstrain bioinformatics pipeline. A dated phylogeny of 5,211 SARS-CoV-2 genomes, along with sampling dates and locations, was retrieved from nextstrain.org/ncov on May 12, 2020. Genomes of non-human origin (n = 13) were discarded from analysis. Polytomies (unresolved divergences represented as a node with >2 descendants) were resolved as branches with an arbitrarily small length of 1 hour, as recommended for adjustment of zero-length risk intervals in Cox regression^47^. Of note, excluding these zero-length branches would bias the analysis by underestimating the number of divergence events in specific regions of the phylogeny. Maximum-likelihood ancestral state reconstruction was used to assign internal nodes of the phylogeny to countries in a probabilistic fashion, taking the tree shape and sampling locations as input data^48^. To prepare data for survival analysis, we decomposed the branches of the dated phylogeny into a set of time-to-event and time-to-censoring intervals (Fig. 1c). Intervals were assigned to the most likely country at the origin of the branch when this country’s likelihood was >0.95. Intervals in which no country reached a likelihood of 0.95 were excluded from further analysis (Extended Data Figs. 1, 2). Finally, intervals during which a change of intervention occurred were split into sub-intervals, such that all covariates, including the country and interventions, were held constant within each sub-interval and only the last subinterval of an internal branch was treated as a time-to-event interval. This interval-splitting approach is consistent with an interpretation of interventions as external time-dependent covariates^18^, which are not dependent on the event under study, namely, the viral divergence event.

### Mixed-effect Cox proportional hazard models

Variations of the divergence rate *λ* in response to non-pharmaceutical interventions were modelled using mixed-effect Cox proportional hazard regression (reviewed in^49^). Models treated the country and phylogenetic branch as random effects to account for non-independence between sub-intervals of the same branch and between branches assigned to the same country. The predictors of interest were not heritable traits of SARS-CoV-2, thus, phylogenetic autocorrelation between intervals was not corrected for. Time-to-event data were visualized using Kaplan-Meier curves with 95% confidence intervals. The regression models had the form

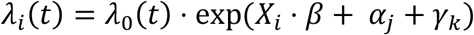

where *λ*_*i*_(*t*) is the hazard function (here, the divergence rate) at time *t* for the *i*th observation, *λ*_0_(*t*) is the baseline hazard function, which is neither specified or explicitly evaluated, *X*_*i*_ is the set of predictors of the *i*th observation (the binary vector of active non-pharmaceutical interventions), *β* is the vector of fixed-effect coefficients, *α*_*j*_ is the random intercept associated with the *j*th phylogenetic branch and *γ*_*k*_ is the random intercept associated with the *k*th country. Country comparison models (Fig. 2d), in which the country was the only predictor and branches were not divided into subintervals, did not include random intercepts. Raw model coefficients (the log-hazard ratios) additively shift the logarithm of the divergence rate *λ*. Exponentiated coefficients exp *β* (the hazard ratios) are multiplicative factors (fold-changes) of the divergence rate. To ease interpretation, hazard ratios were reported as percentage changes of the divergence rate or, equivalently, of the effective reproduction number *R*_*t*_, equal to (exp *β* - 1) × 100. Analyses were conducted using R 3.6.1 (the R Foundation for Statistical Computing, Vienna, Austria) with additional packages *ape, survival* and *coxme*.

### Estimating the effect of combined interventions

Pointwise estimates and confidence intervals of combined interventions were estimated by adding individual coefficients and their variance-covariances. Cox regression coefficients have approximately normal distribution with mean vector *m* and variance-covariance matrix *V*, estimated from the inverse Hessian matrix of the likelihood function evaluated at *m*. From well-known properties of the normal distribution, the distribution of a sum of normal deviates is normal with mean equal to the sum of the means and variance equal to the sum of the variance-covariance matrix of the deviates. Thus, the coefficient corresponding to a sum of coefficients with mean *m* and variance *V* has mean ∑*m* and variance ∑*V*, from which we derive the point estimates and confidence intervals of a combination of predictors. Importantly, summing over the covariances captures the correlation between coefficients when estimating the uncertainty of the combined coefficient.

### Probability of stopping an epidemic

A central question regarding the effectiveness of interventions or combinations thereof is whether their implementation can stop an epidemic by reducing *R*_*t*_ below 1 (Table 2). Suppose that some intervention has an estimated log-hazard ratio 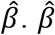 has approximately normal distribution with mean *β* and variance *σ*^2^, written 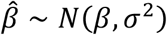. For some fixed value of *R*_0_, the estimated post-intervention reproduction number 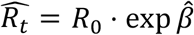. The probability *p* that 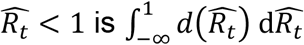 where *d* denotes the probability density function. To solve the integral, remark that 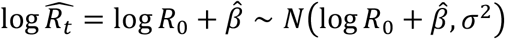. Using a change of variables in the integral and noting that log 1 = 0, we obtain the closed-form solution

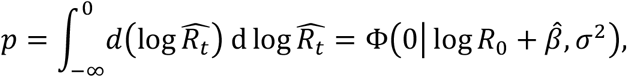

where Ф is the cumulative density function of the normal distribution with mean 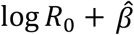 and variance *σ*^2^. By integrating over the coefficient distribution, this method explicitly considers the estimation uncertainty of 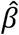 when estimating *p*.

### Potential time-dependent confounders

Time-dependent phylodynamic survival analysis assumes that variations of branch lengths though time directly reflect variations of the divergence rate, which implies that branch lengths are conditionally independent of time given the divergence rate. When the phylogeny is reconstructed from a fraction of the individuals, as is the case in virtually all phylodynamic studies including ours, this conditional independence assumption can be violated. This is because incomplete sampling increases the length of more recent branches relative to older branches^50^, an effect called the diversification slowdown^51,52^. Noteworthy, this effect can be counteracted by a high extinction rate^17,50^, which is expected in our setting and mimicks an acceleration of diversification. Moreover, whether the diversification slowdown should be interpreted as a pure artifact has been controversial^52,53^. Notwithstanding, we considered incomplete sampling as a potential source of bias in our analyses because a diversification slowdown might lead to an overestimation of the effect of non-pharmaceutical interventions. Additionally, the selection procedure used by Nextstrain to collect genomes included in the dated phylogeny possibly amplified the diversification slowdown by using a higher sampling fraction in earlier phases of the epidemic^43^. To verify whether the conclusions of our models were robust to this potential bias, we built an additional multivariable model including the estimated date of each divergence event (the origin of the branch) as a covariate. The possible relation between time and the divergence rate is expectedly non-linear^50^ and coefficient variations resulting from controlling for time were moderate (Extended Data Table 1), thus, we refrained from including a time covariate in the reported regression models as this might lead to overcontrol. Further research is warranted to identify an optimal function of time that might be included as a covariate in phylodynamic survival models to control for sources of diversification slowdown.

### Compartmental epidemiological models

Epidemic dynamics can be described by partitioning a population of size *N* into three compartments, the susceptible hosts *S*, the infected hosts *I*, and the recovered hosts *R*. The infection rate *b* governs the transitions from *S* to *I* and the recovery rate *g* governs the transitions from *I* to *R* (we avoid the standard notation *β* and *γ* for infection and recovery rates to prevent confusion with Cox model parameters). The SIR model describes the transition rates between compartments as a set of differential equations with respect to time *t*,

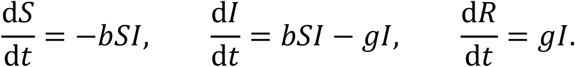

The transition rates of the SIR model define the basic reproduction number of the epidemic, *R*_0_ = *b*/*g*. From a phylodynamic standpoint, if the population dynamics of a pathogen is described as a birth-death model with divergence rate *λ* and extinction rate *μ*, then *R*_*t*_ = *λ*/*μ* or, alternatively, 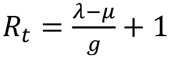^54^. We simulated the epidemiological impact of each individual intervention in SIR models with *R*_0_ = 3 and *g* ^−1^ = 2 weeks based on previous estimates^24,25^, yielding a baseline infection rate *b* = *gR*_0_ = 6. In each model, the effective infection rate changed from *b* to *b* · exp *β* on the implementation date of an intervention with log-hazard ratio *β*. To determine realistic implementation delays, the starting time of the simulation was set at the date of the first local divergence event in each country and the implementation date was set to the observed median delay across countries (see Fig. 3a). All models started with 100 infected individuals at *t* = 0, a value assumed to reflect the number of unobserved cases at the date of the first divergence event, based on the temporality between the divergence events and the reported cases (Extended Data Fig. 3) and on a previous estimate from the U.S. suggesting that the total number of cases might be two orders of magnitude larger than the reported count^55^. Evaluation of the SIR models used the R package *deSolve*.

## Data Availability

Both data and analysis code are available online at https://github.com/rasigadelab/covid-npi.

## Data and software availability

All data and software code used to generate the results are available at github.com/rasigadelab/covid-npi.

## Acknowledgements

We thank Philip Supply, François Vandenesch, Jean-Sébastien Casalegno, Vanessa Escuret, and Christophe Ramière for fruitful discussions and reviews of our work. We thank the GISAID, Nextstrain and OxCGRT teams for making their high-quality datasets available to the community. A list of authors and laboratories contributing SARS-CoV-2 genome sequences is shown in Extended Data Table 3.

## Author contributions

JPR, LJ, TW designed research. JPR, ABarray, JTS, CQ, YV, GD, LJ conducted research. JPR, TW analyzed the data. JPR created figures. JPR, Abal, GD, LJ, PV, BL, TW interpreted the data. All authors wrote the paper.

## Data and material availability

Data and analysis code are available online at https://github.com/rasigadelab/covid-npi.

## EXTENDED DATA

Extended Data Figures 1 to 8 included below

Extended Data Tables 1 and 2 included below

### Other Supplementary Information for this manuscript include the following

Extended Data Table 3. Number of samples, phylogenetic branches, and dates of first detected SARS-CoV-2 local transmission events and non-pharmaceutical interventions in 74 countries. (.xlsx spreadsheet)

Extended Data Table 4. Detailed timeline of implementation and release of non-pharmaceutical interventions against COVID-19 in 74 countries up to May 12, 2020. (.xlsx spreadsheet)

Extended Data Table 5. Authors and laboratories having contributed SARS-CoV-2 genomes included in the dated phylogeny. (.xlsx spreadsheet)

**Extended Data Figure 1.**
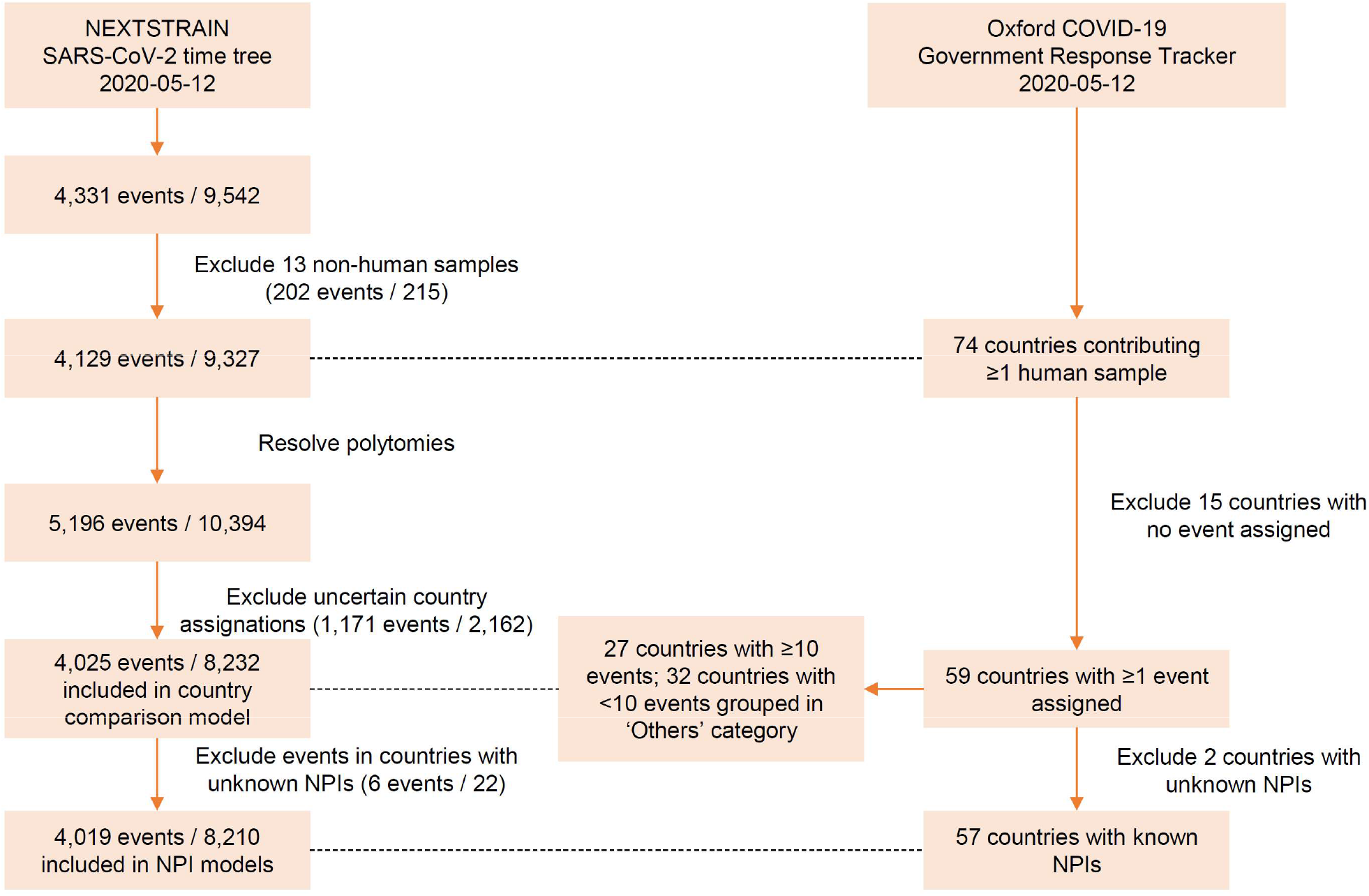
Flowchart of data selection. Events are phylogenetic divergences (tree nodes in the SARS-CoV-2 phylogeny), excluding tree root. Polytomies are unresolved tree nodes representing >1 divergence event. Polytomies were resolved into dichotomies (nodes with exactly 1 divergence) with arbitrarily small interval length. NPI, non-pharmaceutical intervention against COVID-19.

**Extended Data Figure 2.**
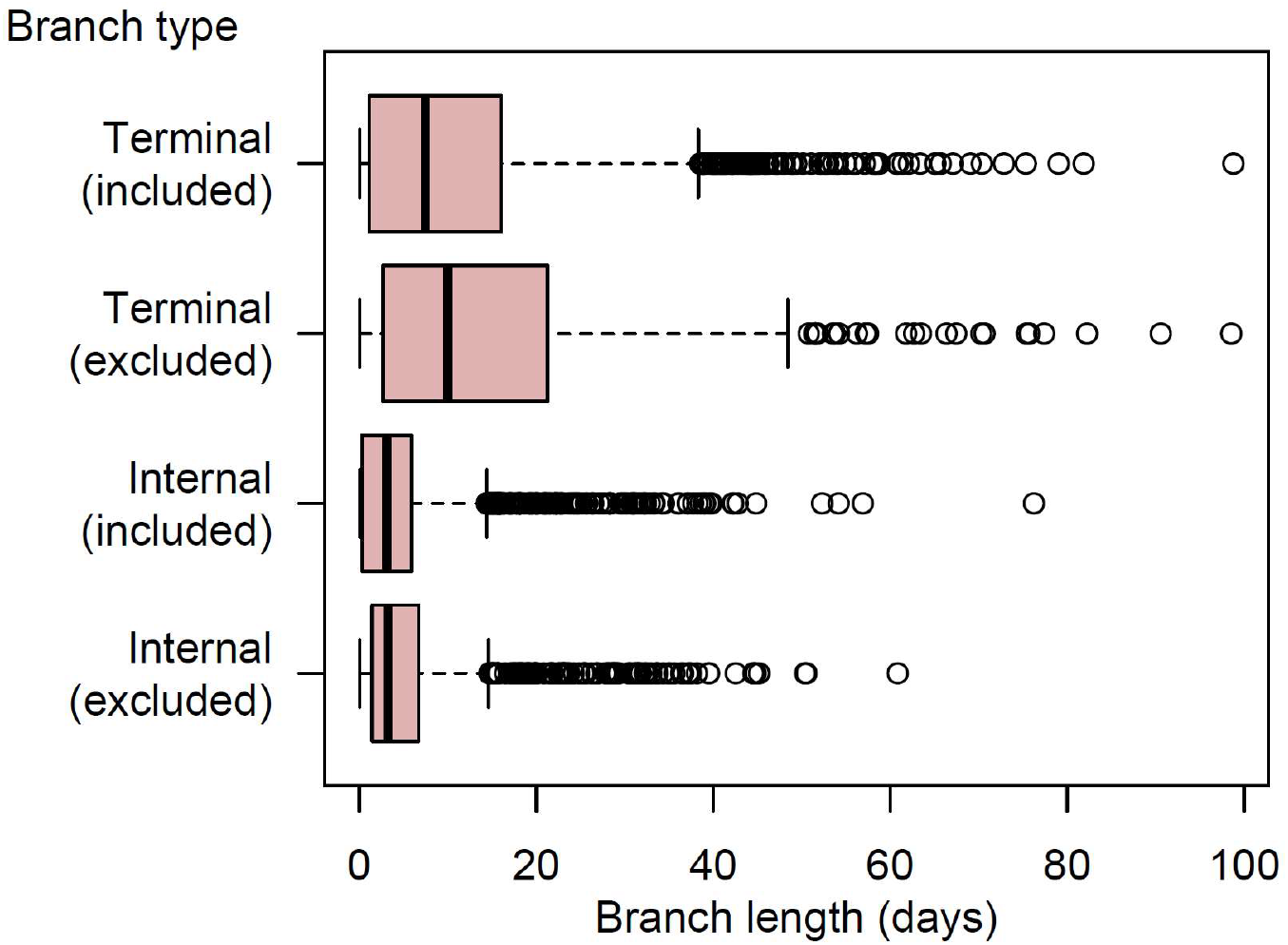
Length distribution in phylogenetic branches with uncertain country assignation. Shown are box-and-whisker plots of the lengths of internal and terminal branches, depending on branch exclusion due to uncertain (<95% confidence) country assignation. Boxes denote interquartile range (IQR) and median, whiskers extend to lengths at most 1.5x the IQR away from the median length, and circle marks denote lengths farther than 1.5 IQR from the median length.

**Extended Data Figure 3.**
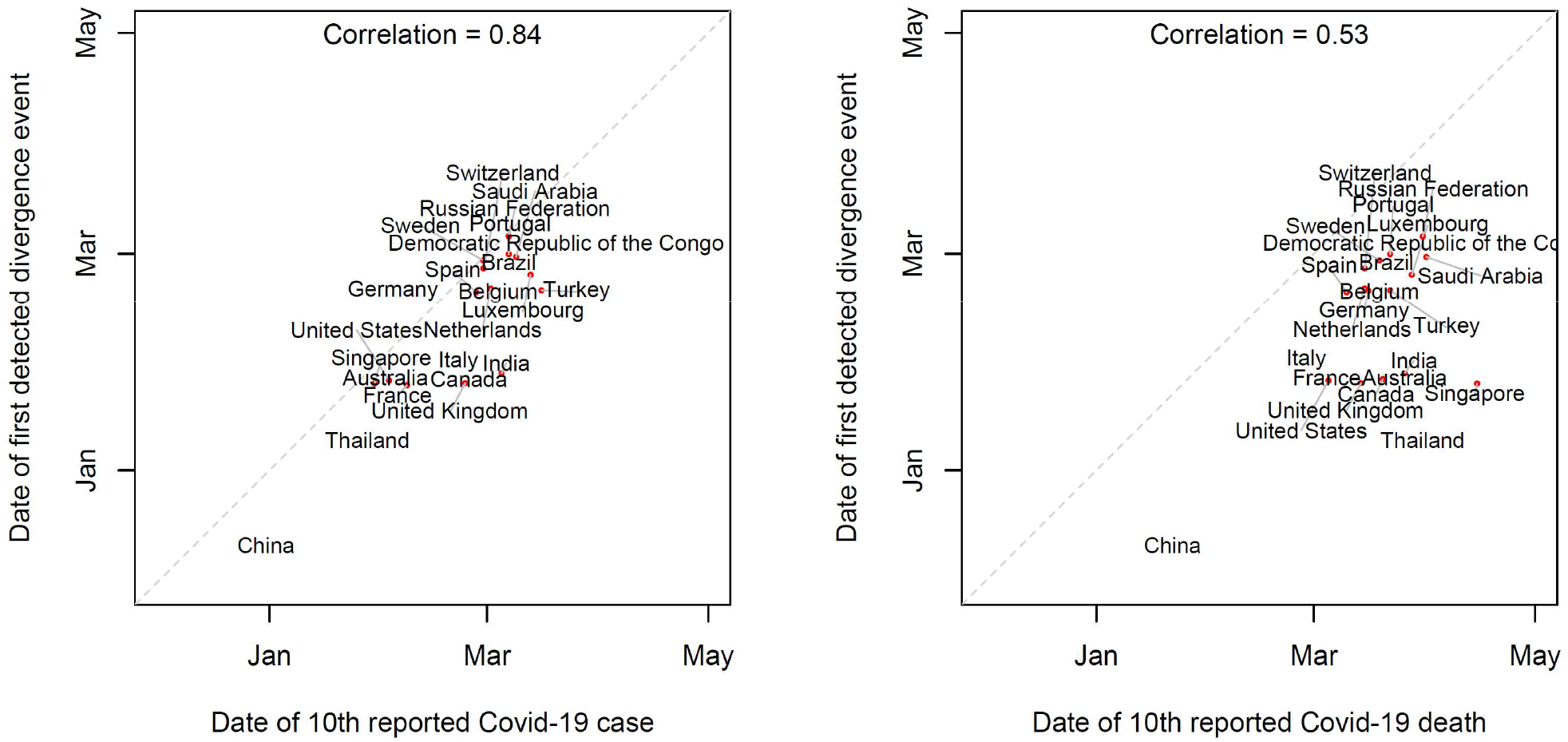
Correlation of reported and estimated epidemic onset dates. Dates of first estimated autochtonous SARS-CoV-2 transmission per country relative to the dates of the 10^th^ reported case (left panel) and the 10^th^ reported death (right panel) in countries with at least 15 assigned internal branches.

**Extended Data Figure 4.**
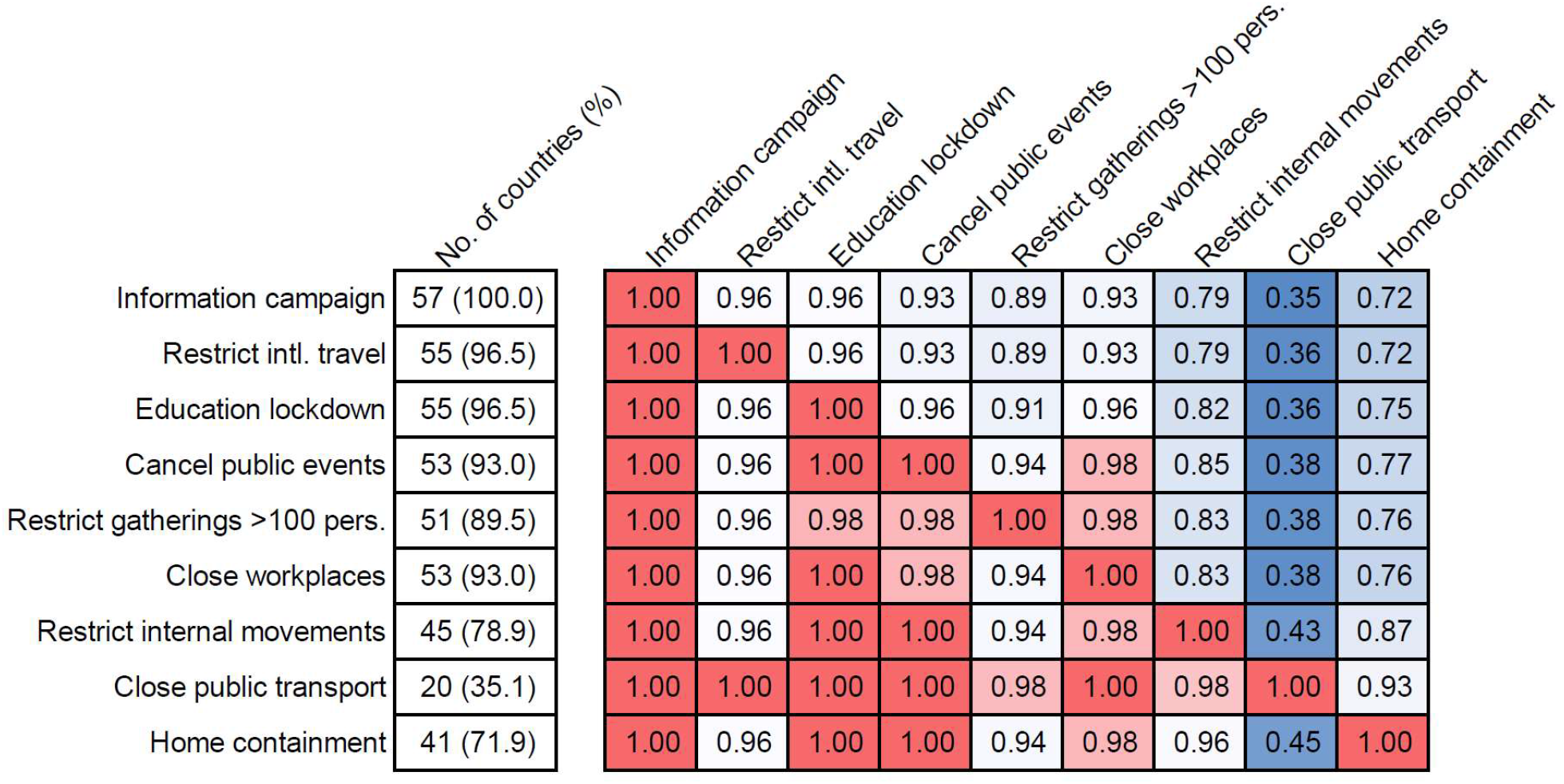
Frequency and timing of implementation of non-pharmaceutical interventions in 57 countries. The first column shows the number and percentage of countries implementing each intervention, independent of other interventions. Matrix cells show the proportion of countries implementing the intervention in column conditional on the implementation of the intervention in row.

**Extended Data Figure 5.**
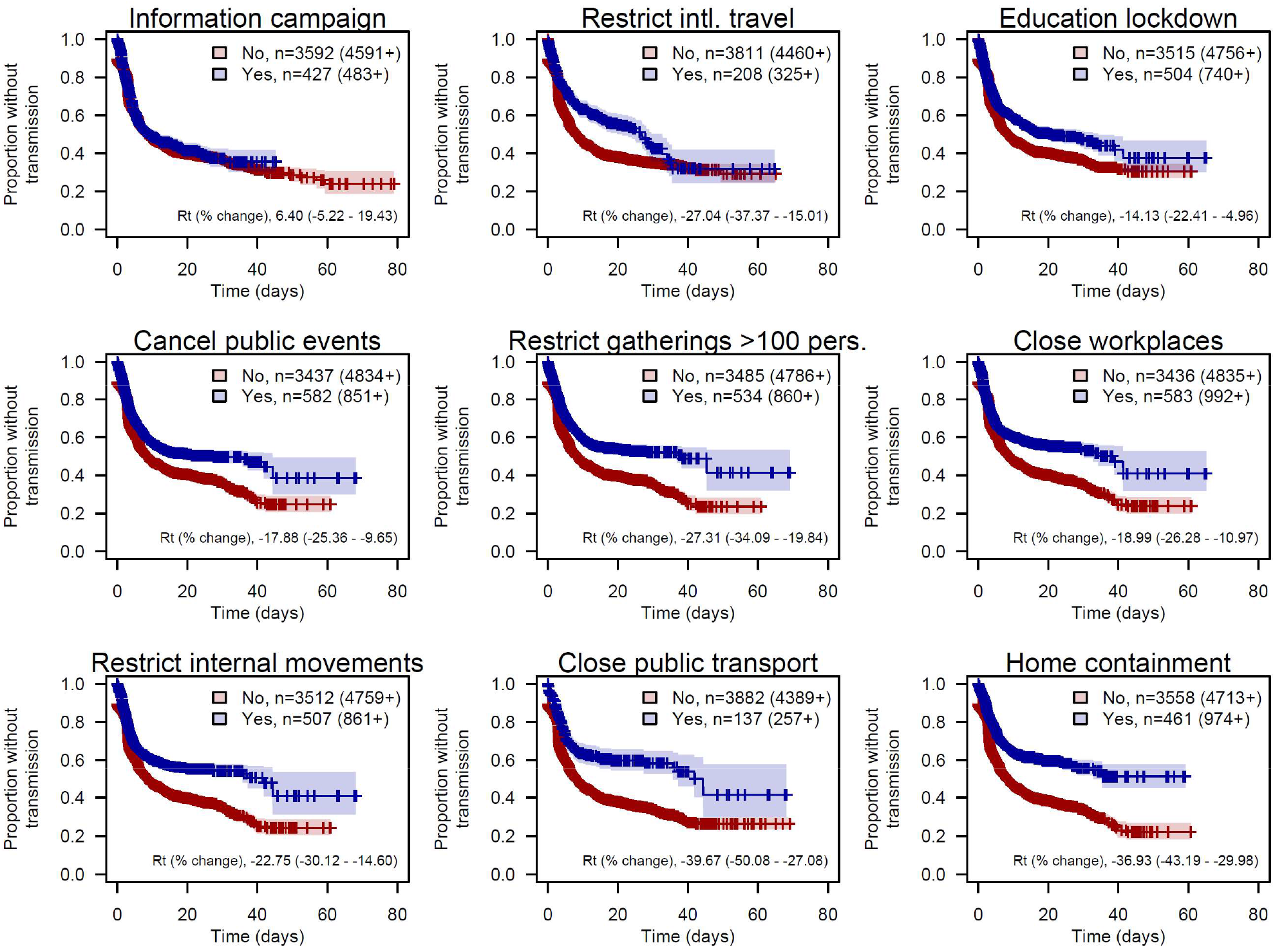
Non-pharmaceutical interventions against COVID-19 correlate with reduced effective reproduction numbers. Data derive from a dated phylogeny of SARS-CoV-2 genomes from 57 countries, with 4,191 internal branches interpreted as time-to-event intervals, and 4,019 terminal branches interpreted as censored intervals, after exclusion of branches with uncertain country assignation. Shown are Kaplan-Meier survival curves of the waiting time without a viral transmission event, stratified on the presence of nine non-pharmaceutical interventions active or not in each country. Sample sizes denote, for each stratum, the no. of time-to-event subintervals (possibly resulting from splitting intervals containing a change of intervention) and, in brackets, the no. of censored subintervals. Percent changes of the effective reproduction number *R*_*t*_ were derived from separate time-dependent mixed-effect Cox regression models treating the country and the branch as random effects.

**Extended Data Figure 6.**
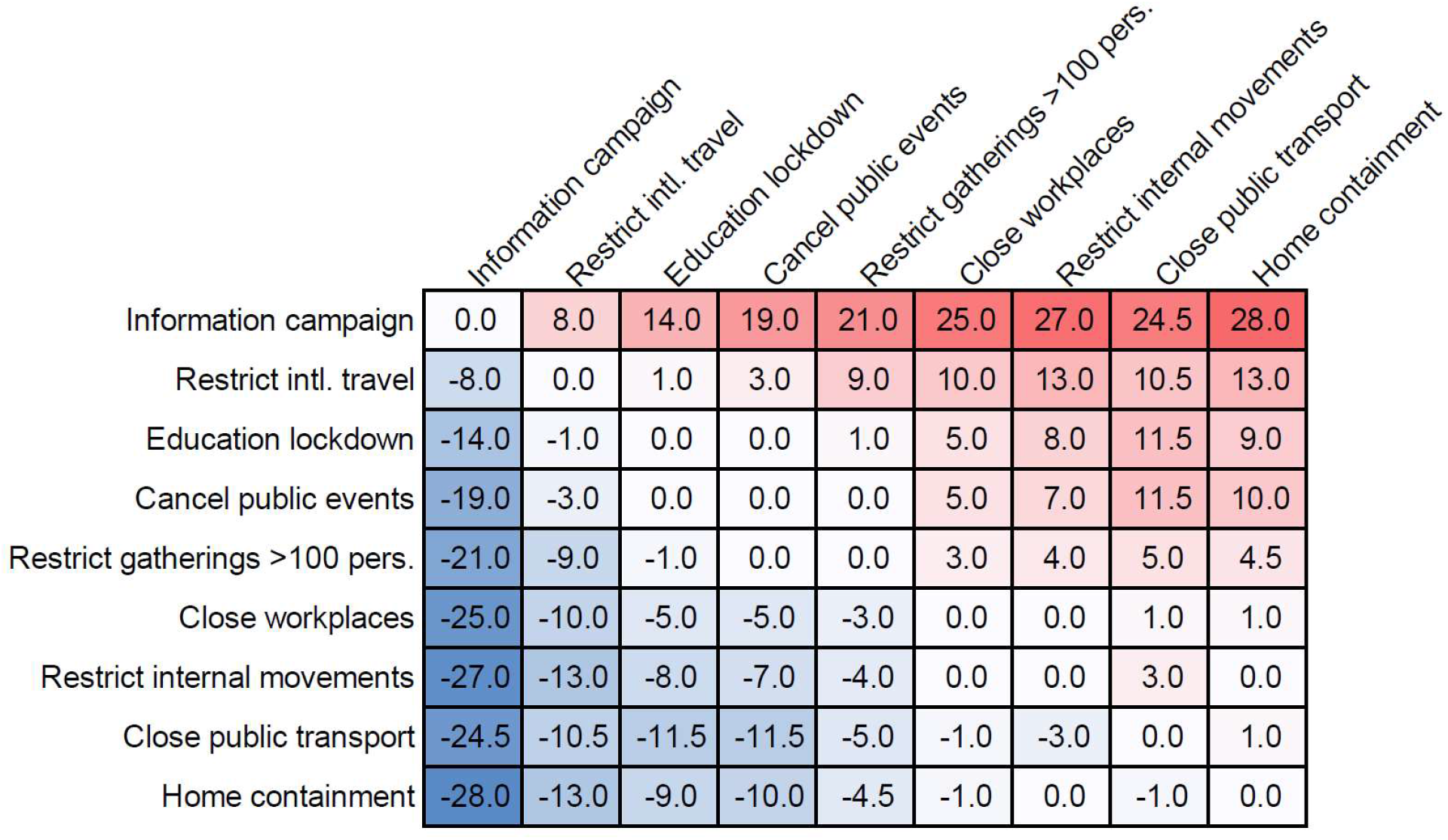
Median delay between implementation of non-pharmaceutical interventions. Shown are the median days elapsed between the implementation of the intervention in the row and that of the intervention in the column, where median is taken across countries that implemented both interventions.

**Extended Data Figure 7.**
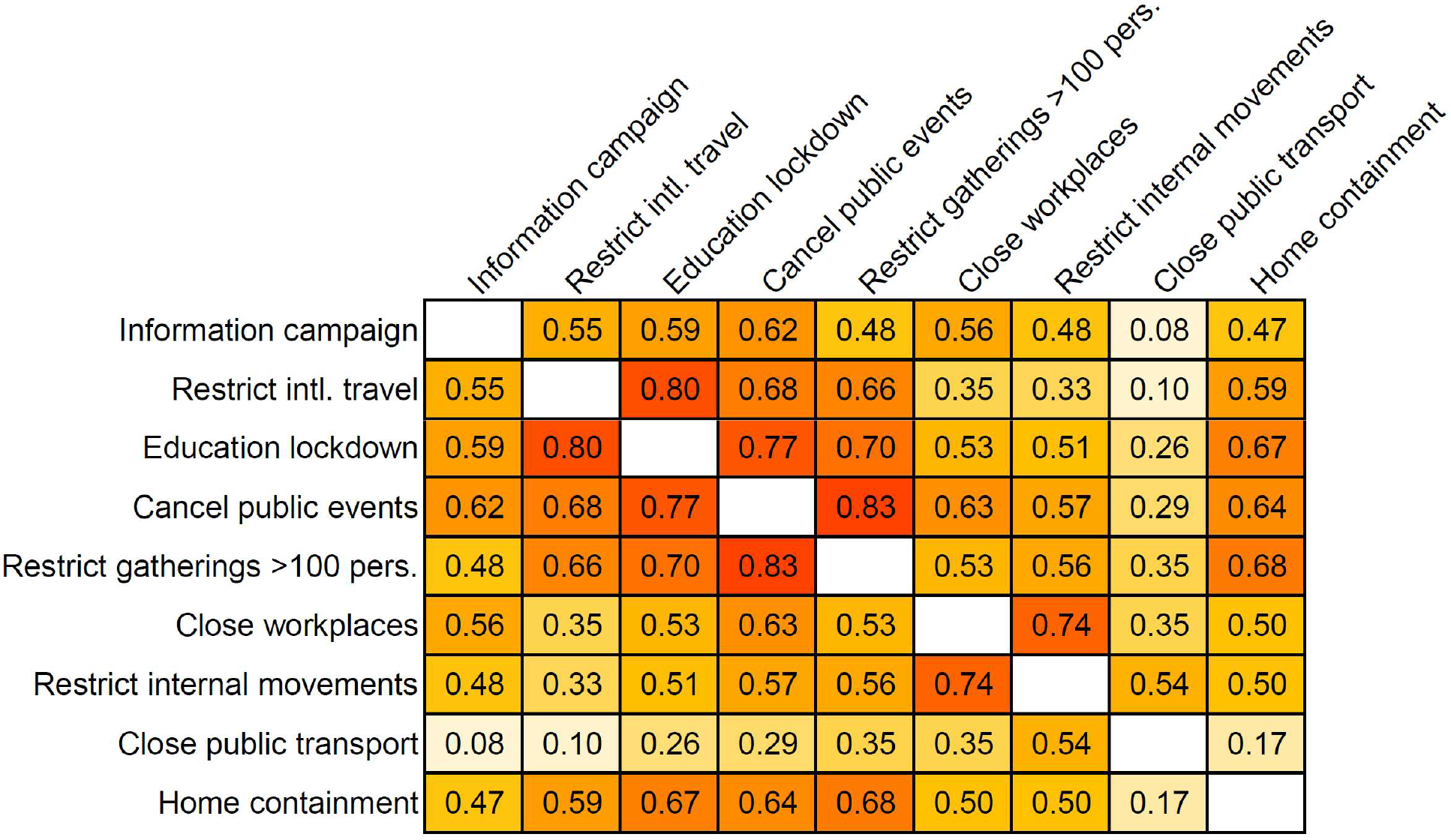
Pearson correlation between non-pharmaceutical interventions. Data derive from 14,829 sub-intervals, including 4,019 time-to-event sub-intervals and 10,810 censored sub-intervals. Sub-intervals result from splitting phylogenetic branches (n = 8,210) in which a change of intervention (activation or release) occurs. Smaller absolute correlations (white) favor the identifiability of intervention effects in multivariable analysis while larger absolute correlations (orange/red) can result into dependencies between model coefficients (see Fig. 3c).

**Extended Data Figure 8.**
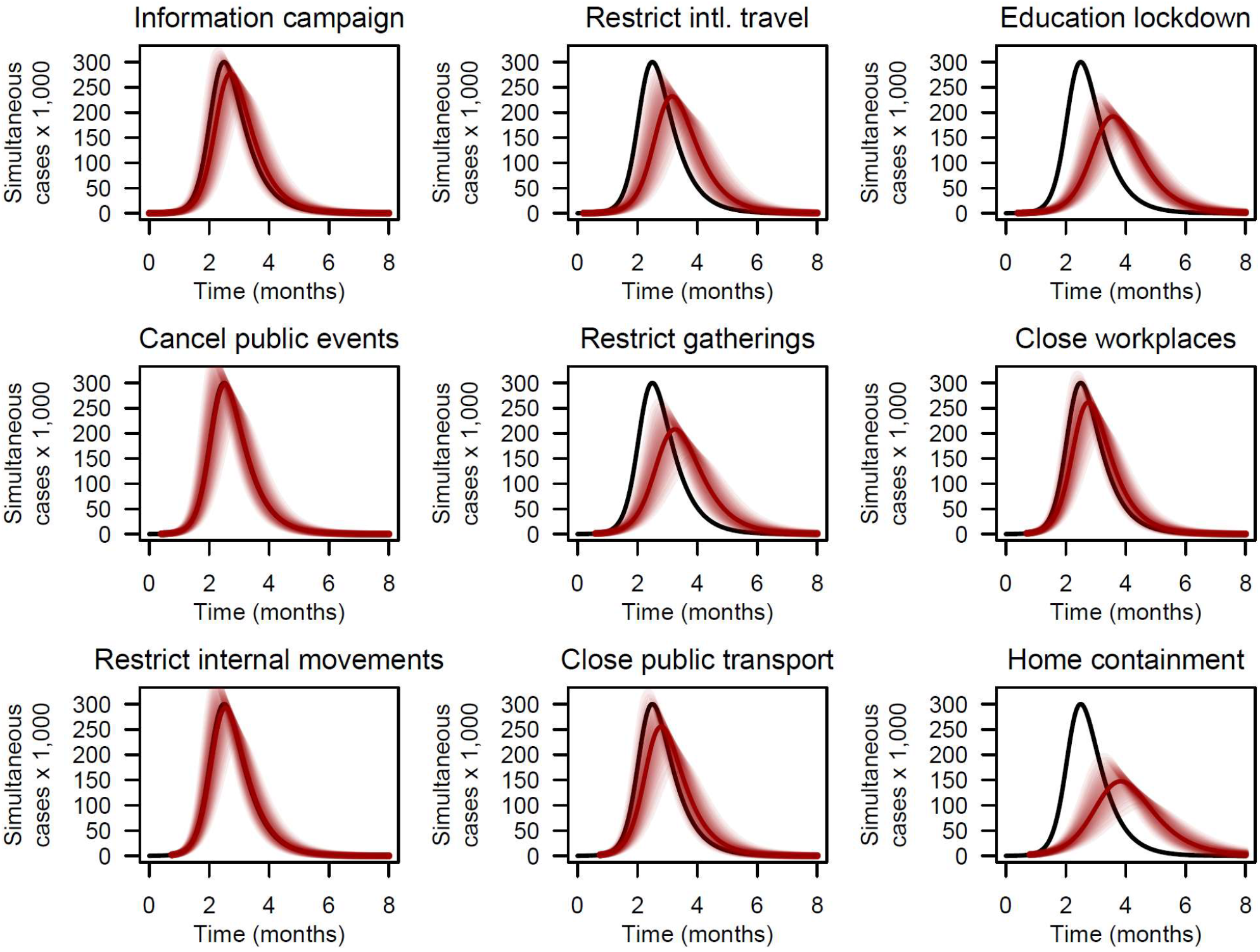
Predicted individual impact of 9 non-pharmaceutical interventions (NPIs) on the number of simultaneous COVID-19 cases in an idealized population of 1 million susceptible individuals. Gray lines represent the case count predicted by an epidemiological SIR model with a basic reproduction number *R*_0_ = 3, as estimated in the absence of NPIs, and a mean infectious period of 2 weeks. For each NPI, the simultaneous case count (red line) and 95% confidence band are derived from an SIR model in which the basic reproduction number is altered as predicted by the mutivariate model coefficients shown in Fig. 3b. The delay between the 100^th^ case and NPI implementation in SIR models coincides with the median delay between the 1^st^ transmission event and the NPI implementation shown in Fig. 3a.

**Extended Data Table 1.** Predicted percent change of COVID-19 effective reproduction number in response to non-pharmaceutical interventions with and without adjustment for time. Data derive from multivariable mixed-effect Cox regression models including one random intercept per country and phylogenetic branch.

| Factor | Relative $R_t$ (% change) | |
| --- | --- | --- |
|  | Base model | Time-adjusted model |
| Elapsed time (per month) | - | -31.4 (-38.9 to -22.9) |
| Information campaign | -6.0 (-17.0 to 6.5) | 3.2 (-9.6 to 17.8) |
| Restrict intl. travel | -16.9 (-27.5 to -4.8) | -11.0 (-22.8 to 2.7) |
| Education lockdown | -25.6 (-33.4 to -16.9) | -21.0 (-29.6 to -11.2) |
| Cancel public events | -1.0 (-14.7 to 15.0) | 1.1 (-13.5 to 18.0) |
| Restrict gatherings >100 pers. | -22.3 (-33.4 to -9.3) | -17.3 (-29.5 to -2.9) |
| Close workplaces | -10.0 (-22.8 to 5.0) | -8.8 (-22.2 to 6.9) |
| Restrict internal movements | -2.2 (-16.8 to 15.0) | 0.8 (-14.8 to 19.2) |
| Close public transport | -11.5 (-26.6 to 6.7) | -9.7 (-25.6 to 9.5) |
| Home containment | -34.6 (-43.2 to -24.7) | -35.4 (-44.0 to -25.4) |

**Extended Data Table 2.** Predicted reduction of the COVID-19 effective reproduction number by non-pharmaceutical interventions implemented alone.

| **Intervention** | **Probability that**<br>**$R_t < 1$ if $R_0 = 1.5$** |
| --- | --- |
| Information campaign | <0.01 |
| Restrict intl. travel | <0.01 |
| Education lockdown | 0.03 |
| Cancel public events | <0.01 |
| Restrict gatherings >100 pers. | 0.03 |
| Close workplaces | <0.01 |
| Restrict internal movements | <0.01 |
| Close public transport | <0.01 |
| Home containment | 0.61 |

NOTE. The probability that $R_t < 1$ was less than 0.01 for all interventions if $R_0 \geq 2$ .

